# Multicenter analysis of COVID-19 hospitalizations and stacking machine learning algorithms for prediction of high-risk patients

**DOI:** 10.1101/2023.06.20.23291685

**Authors:** Reid Shaw, David Bassily, Love Patel, Timothy O’Connor, Robert Rafidi, Perry Formanek

## Abstract

**Objective:** To create and validate an ensemble of machine learning algorithms to accurately predict ICU admission or mortality upon initial presentation to the emergency department.

**Methods:** This is a retrospective cohort study of a multicenter hospital system in the United States. The electronic health record was queried from March 2020 to December 2021 for patients who presented to the emergency department who were subsequently COVID-positive. Associated patient demographics, vitals, and laboratory vitals were obtained. High-risk individuals were defined as those who required ICU admission or died; low-risk individuals did not meet those criteria. The dataset was split into a 3:1 training to testing dataset. A machine learning ensemble stack was built to predict ICU admission and mortality.

**Results:** Of the 3,142 hospital admissions with a COVID positive test, there were 1,128 (36%) individuals labeled as high-risk, and 2,014 (64%) as low-risk. We obtained 147 unique variables. CRP, LDH, procalcitonin, glucose, anion gap, creatinine, age, oxygen saturation, oxygen device, and obtainment of an ABG were chosen. Six machine learning models were then trained over model-specific hyperparameters, and then assessed on the testing dataset, generating an area under the receiver operator curve of 0.751, with a specificity of 95% in predicting high-risk individuals based on an initial emergency department assessment.

**Conclusion:** A novel machine learning model was generated to predict ICU admission and patient mortality from a multicenter hospital system and validated on unseen data.

## 1 Introduction

The coronavirus disease 2019 (COVID-19) pandemic caused by SARS-CoV-2 continues to pose a significant challenge to public health. The clinical outcomes of COVID-19 infection range from asymptomatic to serious complications and death. While vaccinations have decreased rates of transmission and treatments have improved rates of mortality, the pandemic remains an ongoing challenge^1–3^. It is essential for effective screening and triaging of patients with COVID-19 as this can mitigate the burden on the healthcare system and aid in the allocation of resources to those in greatest need^4^.

One method used to triage high-risk individuals is applying a scoring system that predicts mortality and a higher level of care needs. Currently, multiple scoring systems are used nationwide. These include National Early Warning Score (NEWS)^5^, Modified Early Warning Score (MEWS)^6^, Between the Flags (BTF)^7^, Quick Sequential Sepsis-Related Organ Failure Assessment (qSOFA)^8^, and Systemic Inflammatory Response Syndrome (SIRS)^9^. However, these scoring systems were not specifically created for COVID infections and do not account for known predictors of COVID-related mortality. To meet these needs, there have been multiple groups creating new scoring systems with more advanced statistical methods. For example, individual machine learning (ML) algorithms have been applied to predict mortality in patients with COVID-19^10–12^. However, stacking ML algorithms into an ensemble is an effective method in increasing model performance over a variety of clinical and non-clinical problems^13–15^.

In this study, we sought to develop a ML ensemble that predicts high and low-risk patients who have tested positive for COVID based on a set of variables commonly obtained upon initial presentation to the emergency department.

## 2 Methods

### 2.1 Study design and data collection

This is an IRB-approved, retrospective cohort study from three academic hospitals in the greater Chicago area – Loyola University Medical Center, Gottlieb Memorial Hospital, and MacNeal Hospital. The electronic health record (EHR) software (Epic Systems; Verona, WI) was queried from January 1, 2020, to December 31, 2021. Inclusion criteria included all individuals admitted to one of the three hospitals who tested positive for COVID-19 via polymerase chain reaction (PCR). For each patient, lab values, demographics, and disposition were collected. Common lab values include complete metabolic panel (CMP), complete blood count (CBC) with differential, magnesium, phosphorus, procalcitonin, d-dimer, lactate dehydrogenase (LDH), c-reactive protein (CRP), troponin, brain natriuretic peptide (BNP), ferritin, international normalized ratio (INR), activated partial thromboplastin time (aPTT), and arterial blood gas (ABG). Vital signs include temperature, systolic and diastolic blood pressure, heart rate, oxygen saturation, respiratory rate, and first oxygen device. Vitals used in model generation were those that were first recorded upon presentation to the hospital. Comorbidities were those that are included in the Charlson comorbidity index, a composite score used to predict 10-year survival^16^. Other variables include race, gender, ethnicity, height, weight, and body mass index (BMI).

We defined high-risk patients as those that were discharged to hospice, died in the hospital, and/or were admitted to the intensive care unit (ICU). Low-risk patients were defined as those that do not meet any of the high-risk requisites. The complete patient cohort was pseudo-randomly split into a 3:1 training to testing dataset. The training dataset was further partitioned into five repeats of v-fold cross validation resamples for robust model evaluation. Importantly, all of the feature engineering and feature selection were performed on the training dataset. The testing dataset was used only for final model evaluation.

### 2.2 Feature selection and engineering

Feature engineering was used to reformat the set of predictors to improve model performance. The neutrophil to lymphocyte ratio (NLR) was calculated from the CBC. Variables to indicate the presence of high-risk lab tests were created, including troponin, BNP, or ABG. Nominal predictors with infrequently occurring (<10%) values were grouped into an “other” category. Nominal variables were encoded into dummy variables, a numeric binary model term for the number of levels of the original data. Terms with zero variance were eliminated. Missing data were imputed with k-nearest-neighbors. Features with zero variance or Spearman correlation values above 0.9 were eliminated.

Feature selection was used to narrow the list of predictors to a set that improve model performance. Four unique methods of feature selection were applied before model generation: domain expertise, univariate analysis, stepwise logistic regression, and an embedded random forest model. The statistical significance of differences for each predictor was analyzed using a two-sided t-test. Stepwise logistic regression models were generated on the training dataset across the bootstrap resampled training data. This method iteratively eliminated each variable to assess the change in model performance. Three distinct methods of data imputation were assessed within an elastic net model: k-nearest neighbors, bagged trees, and mean imputation. The elastic net models were tuned over a variety of L1 and L2 regularization values. The random forest model was tuned across model-specific hyperparameters across each training resample.

### 2.3 Ensemble stacking

We assessed six distinct supervised ML classifiers. The classifiers include random forest (RF), flexible discriminant analysis (FDA)^17^, radial basis function supervised vector machine (SVM), naïve Bayes (NB), elastic net (EN), and extreme gradient boosted trees (XGBoost)^18^. Each classifier was tuned over a Latin hypercube of model-specific hyperparameters across the resampled training dataset. All model predictions were then combined into a new data frame and least absolute shrinkage and selection operator (LASSO) was applied as meta learner across six distinct penalties. A higher penalty generally results in fewer members being included in the final model stack. Ultimately, this process determines the stacking coefficients of the individual models and selects those with non-zero stacking coefficients for the final ensemble stack.

### 2.4 Software

All statistical analysis was performed with R programming language v.4.0.3^19^. Packages used were ‘tidymodels,’^20^ ‘tidyverse,’^21^ ‘stacks,’^22^ ‘workflowsets,’^23^ ‘pROC,’^24^ ‘readxl,’^25^ ‘RColorBrewer,’^26^ ‘viridis,’^27^ ‘GGally,’^28^ and ‘corrplot.’^29^

## 3 Results

### 3.1 Cohort characteristics

COVID-19-positive PCR results ranged from March 17, 2020 to December 13, 2021 (**Figure 1A**). In total, there were 4,685 unique admissions with a COVID positive test. Those admissions consisted of 3,995 unique patients -- of which, 3,142 patients had labs and a COVID positive test within three days of admission. 147 unique variables were queried from the EHR. The complete patient cohort was then split, creating a training and testing dataset of 2,356 and 786 individuals, respectively (**Supp. Table 1**) The patient ages ranged from 0 to 104 years with a median age of 60. 1,483 (47%) patients were women. 1644 (52%) were White, 812 (26%) were Black, and 602 (19%) identified as another race (**Table 1**). The median Charlson comorbidity index was 3, with a minimum of 0 and a maximum of 23 (Table 1). The majority of individuals were discharged home (**Supp. Table 1**). 1,148 (36%) patients were labeled as high-risk and 2,014 (64%) were labeled as low-risk (**Table 1**).

**Table 1:** Overview of the training and testing dataset.

|  | Train (N = 2,356) | Test (N = 786) | Cohort (N = 3,142) |
| --- | --- | --- | --- |
| Risk |  |  |  |
| High | 846 | 282 | 1,128 |
| Low | 1510 | 504 | 2,014 |
| Gender |  |  |  |
| Male | 1258 | 416 | 1,674 |
| Female | 1098 | 370 | 1,468 |
| Race |  |  |  |
| White | 1212 | 414 | 1,626 |
| Black | 597 | 211 | 808 |
| Asian | 43 | 11 | 54 |
| Other | 464 | 136 | 600 |
| First O2 device |  |  |  |
| Room Air | 1880 | 623 | 2,503 |
| Nasal cannula | 246 | 100 | 346 |
| Mask | 109 | 29 | 138 |
| Endotracheal tube | 20 | 10 | 30 |
| Vitals |  |  |  |
| Age | 58.7 (0 - 104) | 58.3 (0 - 101) | 58.6 (0 - 104) |
| Heart rate | 97 (0 - 288) | 96 (0 - 192) | 97 (0 - 288) |
| Systolic blood pressure | 131 (53 - 255) | 131 (79 - 241) | 131 (53 - 255) |
| Diastolic blood pressure | 80 (11 - 166) | 75 (35 - 254) | 75 (11 - 254) |
| Respiratory rate | 22 (8 - 109) | 22 (9 - 91) | 22 (8 - 109) |
| Oxygen saturation | 94 (30 - 100) | 94 (50 - 100) | 94 (30 - 100) |
| Temperature | 99.1 (82.4 - 105.8) | 99.1 (92 - 104) | 99 (82.4 - 105.8) |
| Body mass index | 30.8 (11 - 88) | 30.8 (11 - 89) | 30.8 (11 - 88) |
| Labs |  |  |  |
| Anion gap | 12.2 (1 - 35) | 12.2 (4 - 41) | 12.2 (1 - 41) |
| C-reactive protein | 77 (0 - 431) | 80 (0.6 - 469) | 78 (0 - 469) |
| Creatinine | 1.58 (0.19 - 32.5) | 1.55 (0.19 - 16.8) | 1.57 (0.19 - 32.5) |
| Glucose | 156 (20 - 1355) | 160 (42 - 896) | 157 (20 - 1355) |
| Lactate dehydrogenase | 377 (108 - 3191) | 368 (88 - 2550) | 374 (88 - 3191) |
| Procalcitonin | 1.3 (0.02 - 97.1) | 0.78 (0.02 - 52.2) | 1.17 (0.02 - 97.1) |
| Comorbidities |  |  |  |
| Charlson score | 4 (0 - 23) | 4 (0 - 20) | 4 (0 - 23) |

**Figure 1.**
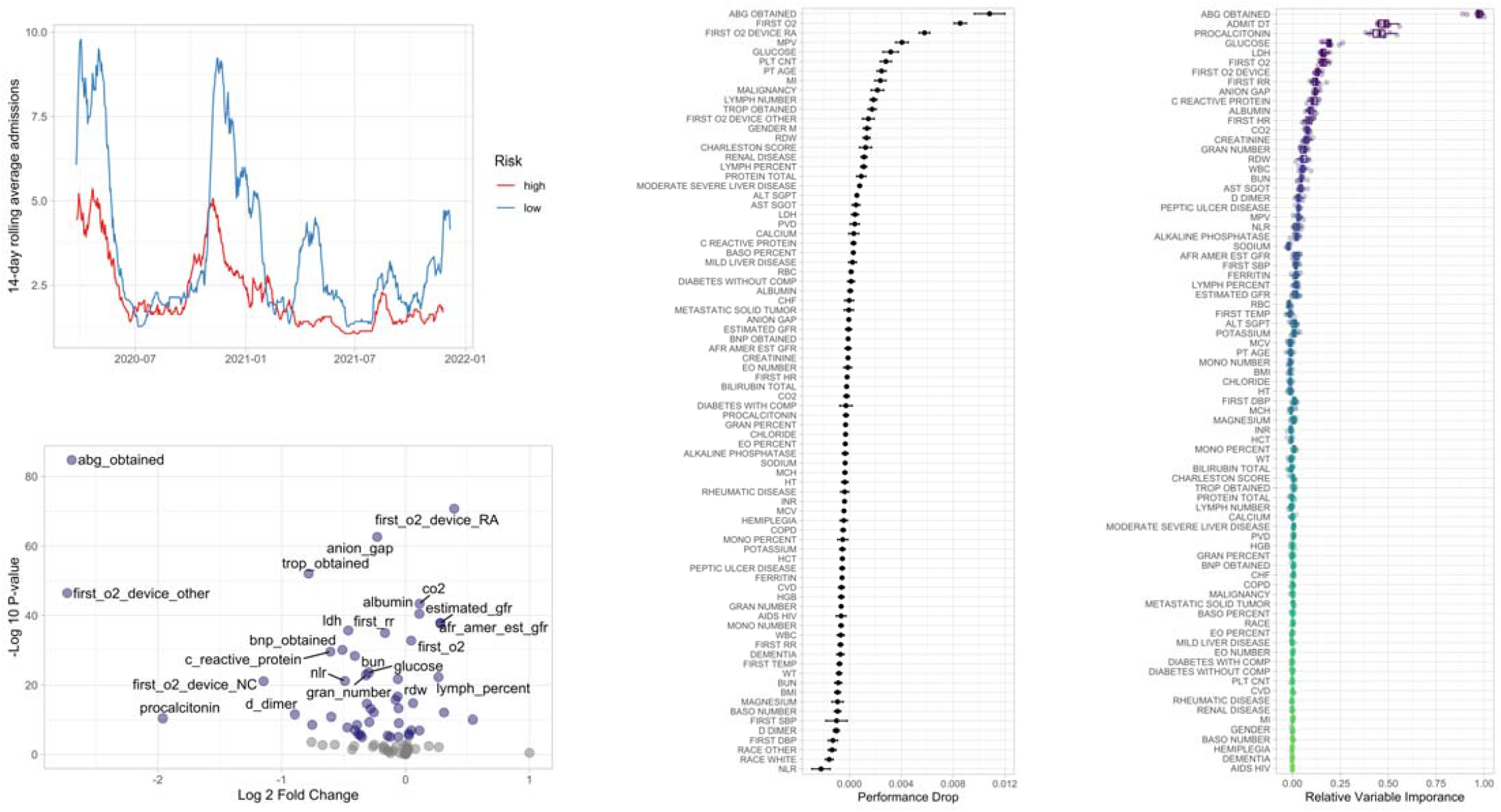
A)14 day rolling average of COVID hospitalizations split into high and low-risk from January 1, 2020 to December 31, 2021. B) Volcano plot of the two-sided t-test of numerical variables. Statistically significant variables are highlighted in purple. A negative log 2 fold change is indicative of variables associated with high-risk patients. C) Iterative elimination of variables within multiple logistic regression models to assess variable importance. Positive performance drop indicates worse model performance with the elimination of that variable. D) Relative variable importance calculated by Gini impurity within 10 random forest models. Higher values indicate greater variable importance within the model.

### 3.2 Feature selection and engineering

Sixty-one variables were statistically different from the high and low-risk groups (**Figure 1B**). The three most significant variables were whether an ABG was obtained, if the patient was on room air, and the degree of elevation of the anion gap (**Figure 1B**). The largest fold change among laboratory tests was procalcitonin; with an average value of 2 among high-risk patients and an average of 0.57 in low-risk patients (**Figure 1B**). Initial presentation of patients on room air, high GFR, and high lymphocyte percentage is predictive of low-risk (**Figure 1B**). There was no significant difference in model performance when missing variables were imputed across different statistical techniques (**Supp. Figure 1**). 82 predictors were assessed in the stepwise LR models, generating 82 unique models. Average model performance decreased most with the elimination of whether an ABG was obtained, initial presenting oxygen saturation, breathing on room air, mean platelet value, glucose, platelet count, and patient age (**Figure 1C**).

The RF model was tuned across 25 model-specific hyperparameters, including the number of predictors at each split, the number of trees, and the minimum number of data points in a node that are required for further splitting. These hyperparameters were tuned over 10 bootstrap resamples of the training dataset. In general, model performance improved with an increasing number of trees, low number of predictors at each split, and a low node size (**Supp. Figure 2B**). The RF model with the greatest AUROC consisted of 986 trees, 14 predictors at each split, and a node size of 3. This model was then pseudo-randomly fit 10 times to calculate Gini Impurity of each variable, a measure of variable importance. The variables with the highest importance were whether an ABG was obtained, the admit date, procalcitonin, glucose, LDH, and the first O2 device (**Figure 1D**).

**Figure 2.**
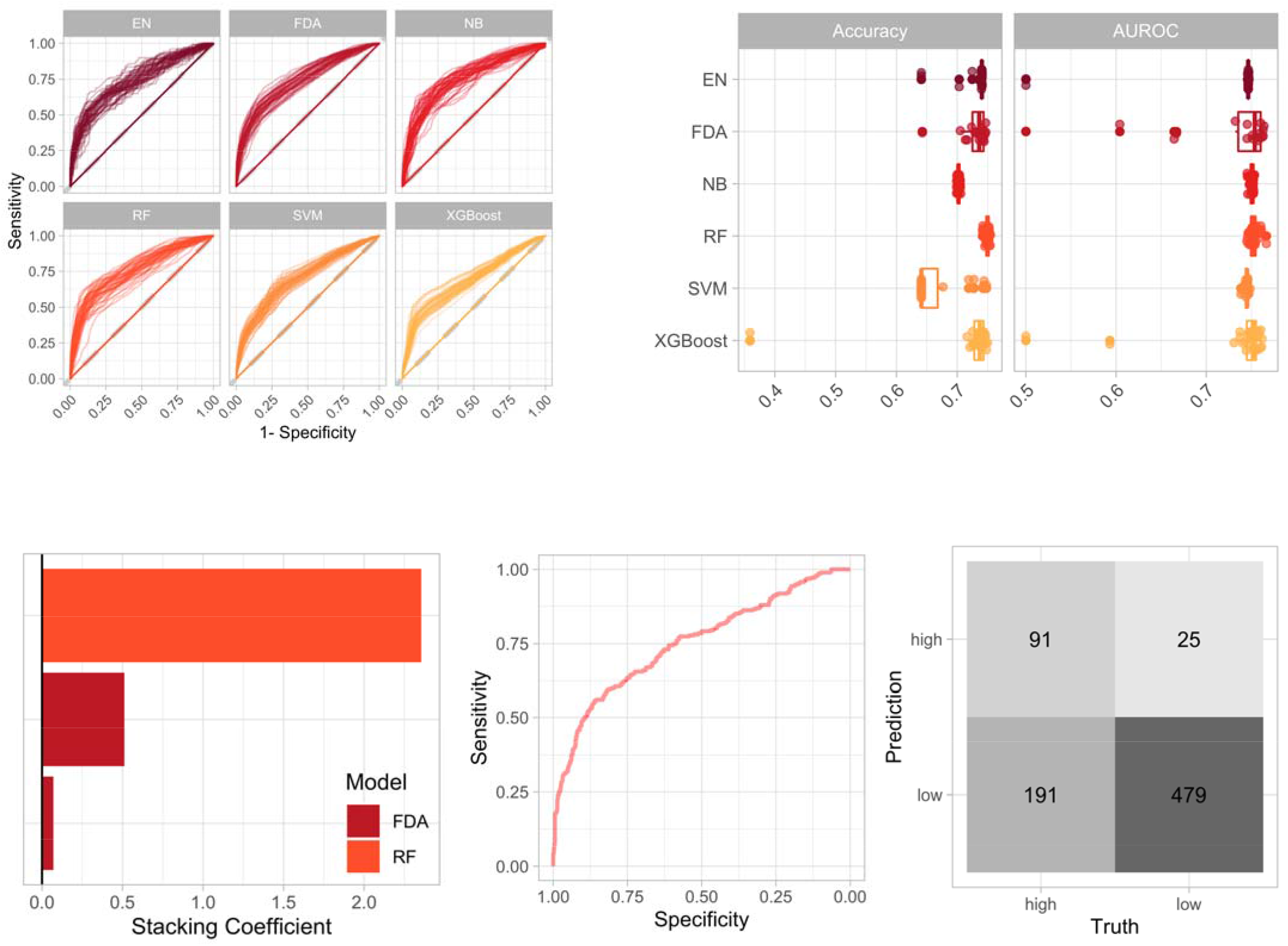
A) AUROC of the six candidate machine learning classifiers across the resampled training dataset. B) Accuracy and AUROC of the six candidate members across the training dataset. C) Stacking coefficient of the three candidate members that compose the ensemble stack. D) AUROC of the ensemble stack when assess on the testing dataset. E) Confusion matrix of the ensemble stack when assessed on the testing dataset.

Based on the orthogonal feature selection methods, the final variables chosen for ensemble stacking were CRP, LDH, procalcitonin, glucose, anion gap, creatinine, age, oxygen saturation, oxygen device, and obtainment of an ABG.

### 3.2 Ensemble stacking

Each model was tuned across 30 model-specific hyperparameters using a Latin hypercube, generating 1,971,960 predictions (**Figure 2A**). The maximum AUROC for EN was 0.747, 0.764 for FDA, 0.753 for NB, 0.767 for RF, 0.749 for SVM, and 0.762 for XGBoost (**Figure 2B**). The maximum accuracy for EN was 0.741, 0.747 for FDA, 0.706 for NB, 0.755 for RF, 0.748 for SVM, and 0.750 for XGBoost (**Figure 2B**). Out of 166 possible candidate members, the ensemble retained one RF classifier and two FDA classifiers with a LASSO penalty of 0.1 (**Figure 2C**). The ensemble was then fit to the testing dataset and achieved a final AUROC of 0.751 with a specificity of 95% at a sensitivity of 32%, generating a NPV of 72% and a PPV of 78% (**Figure 2D, E**).

## 4 Discussion

In this study, we developed an ensemble stack of ML algorithms to identify high-risk individuals initial presentation to the hospital who tested positive for COVID-19 via PCR. We queried the electronic health records of three Chicago-area hospitals, identifying 3,142 cases for analysis. These patients were labeled as high and low-risk based on whether they were discharged to hospice, died in the hospital, and/or admitted to the ICU. We used this definition as it is likely to be most clinically relevant. This stratification potentially allows for the early identification of patients at the greatest risk and the subsequent allocation of resources to those in greatest need. We applied six distinct ML algorithms, combining them into an ensemble stack used to make final predictions on the testing dataset. To our knowledge, this is the first time an ensemble stack has been used in the analysis of COVID-19. An ensemble stack has many benefits over traditional ML methods as it combines the strengths of the individual models while minimizing each model’s limitations, often outperforming the individual classifiers^13,14^. The final ensemble stack had an AUROC of 0.75 on the testing dataset – comparable to the training results, thus decreasing the likelihood of model overfitting. Our model demonstrated a 95% specificity at a sensitivity of 32%. While the NPV and PPV will vary across time as prevalence of high and low-risk individuals change, the current NPV and PPV were 71% and 78%, respectively. These results demonstrate an initial proof-of-concept in using an ensemble stack to generate prognostic scoring systems in COVID-19.

Feature selection and engineering are an integral part of the generation of any ML algorithm. This process reduces the computational training time, may improve model performance, and, in this case, limit the number of input variables to make a clinically relevant tool. In this study, we utilized an orthogonal approach to select features. Domain expertise and literature review were utilized to select an initial set of variables to query within the EHR. Next, we performed a univariate statistical analysis, and generated a logistic regression wrapper and embedded RF model. BMI, a variable previously identified as a predictor for severe COVID-19, was not identified as a predictive variable through the wrapper or embedded model^30^. This is possibly due to the ability of ML models to identify trends in data that are not seen in more conventional statistics^31^. Although the admission date is a clear predictor of risk in many of our analyses, it was not chosen as a final feature because it may change with new COVID variants and medicines^32^. The obtainment of an ABG was the most predictive variable through multiple analysis methods. However, this is a subjective decision made by the provider that may vary depending on culture and across institutions. We included this variable in the final model, but it may limit broader applicability. Other predictors within the ensemble include the laboratory markers CRP, procalcitonin, glucose, anion gap, creatinine, and LDH. CRP, an acute phase reactant, has previously been shown to be associated with COVID-19 severity and used in other ML COVID-19 prognostic models^33,34^. LDH, an important enzyme used in the anaerobic metabolic pathway, is also associated with COVID-19 severity^35^. Similarly, multiple studies have identified kidney injury as common in severe COVID-19^36^. Non-lab values included in the ensemble stack are oxygen saturation at presentation, initial oxygen device, and patient age – all of which are strongly associated with COVID-19 severity^37^.

There are several limitations to this study. First, the hospitals queried in this study are located in the greater Chicago area, and thus may not represent results from distinct geographical regions across the United States or in other countries. Additionally, there was no external validation to assess model performance. Vaccination status, which is a strong predictor of disease severity,^38^ was not included as this variable was incompletely captured in many of the hospital records.

In comparison to other prognostic scoring systems, we looked at one point in time to make risk predictions, whereas other prognostic scoring systems continually re-evaluate patients throughout hospitalization. Including variables from different time points throughout hospitalization may have led to improved model metrics. The last limitation of this study is that we are unable to show that identifying high-risk individuals translates into improved patient outcomes^39^. As health care continues its transition into utilizing big data for clinical decision-making, it will remain important to show that predictive analytics improve outcome^40^. Importantly, this ML approach is easy to deploy, and retrain as more data becomes available which will improve predictive performance.

## Data Availability

All data are not currently available.

## Acknowledgements

Thank you to Susan Zelisko for graciously querying data from the EHR.

## Author Contributions

Conceptualization: R.S.; methodology: R.S.; formal analysis: R.S., P.F., D.B., T.O., R.R.; writing - original draft: R.S.; writing - review and editing: R.S., P.F. D.B., T.O., R.R. All authors have read and agreed to the published version of the manuscript.

## IRB statement and informed consent

The study was reviewed by the Loyola University Medical Center IRB (study ID: 215607), determining that the study meets federal and university criteria for exemption. All data in the data repositories are de-identified.

## Abbreviations

ABG: arterial blood gas
aPTT: activated partial thromboplastin time
AUROC: area under the receiver operating characteristic curve
BMI: body mass index
BNP: brain natriuretic peptide
BTF: between the flags
CBC: complete blood count
CMP: complete metabolic panel
COVID-19: coronavirus disease 2019
CRP: c-reactive protein
EHR: electronic health record
EN: elastic net
FDA: flexible discriminant analysis
GFR: glomerular filtration rate
ICU: intensive care unit
INR: international normalized ratio
LASSO: least absolute shrinkage and selection operator
LDH: lactate dehydrogenase
ML: machine learning
MEWS: modified early warning score
NB: naïve bayes
NEWS: national early warning score
NLR: neutrophil to lymphocyte ratio
NPV: negative predictive value
PCR: polymerase chain reaction
PPV: positive predictive value
qSOFA: quick sequential organ failure assessment
RF: random forest
SIRS: systemic inflammatory response syndrome
SVM: supervise vector machine
XGBoost: extreme gradient boosted trees.

## Figures

**Supplementary Figure 1.**
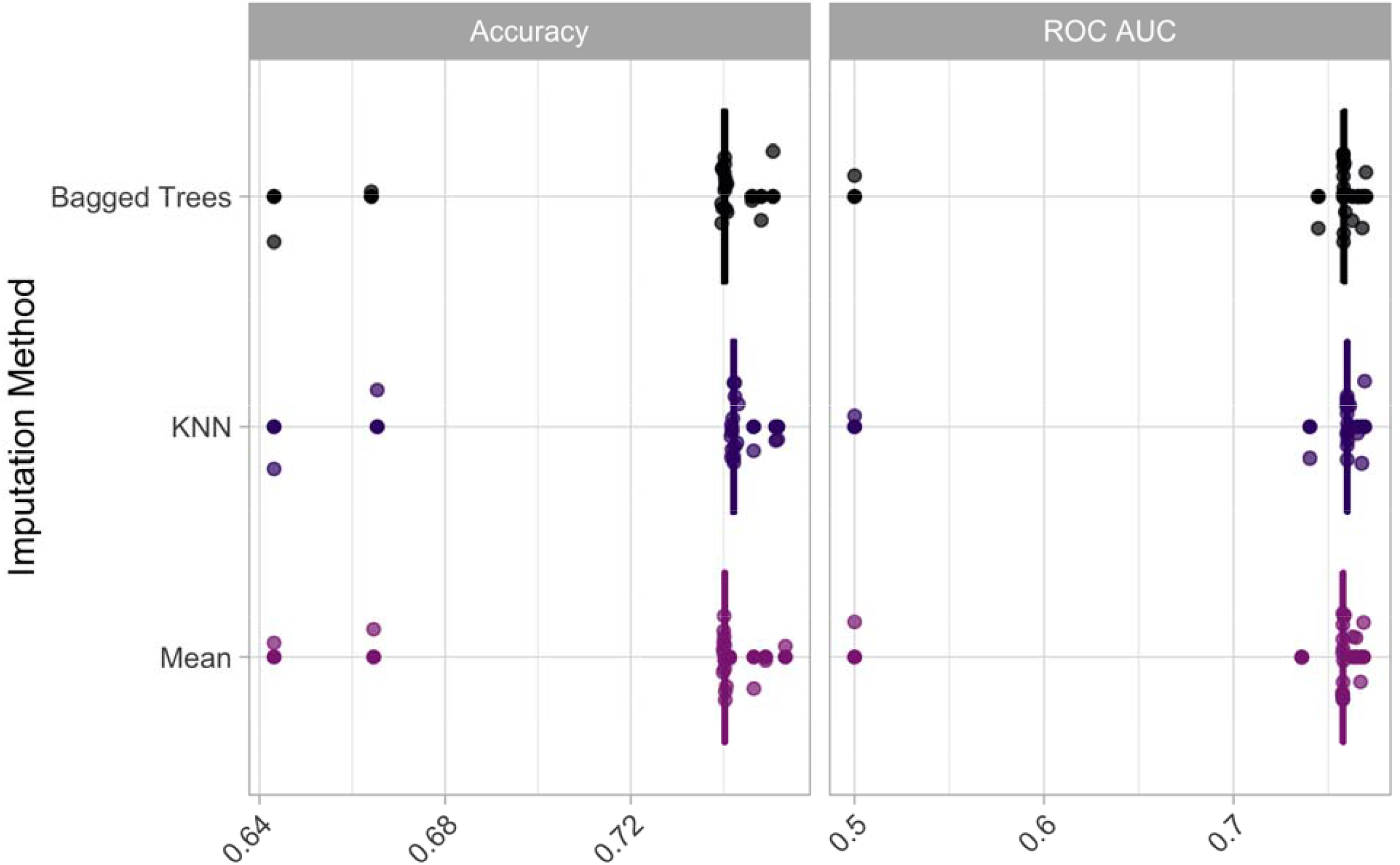
Accuracy and AUROC of the three imputation methods assessed in the training dataset: bagged trees, K-nearest neighbors, and mean imputation.

**Supplementary Figure 2.**
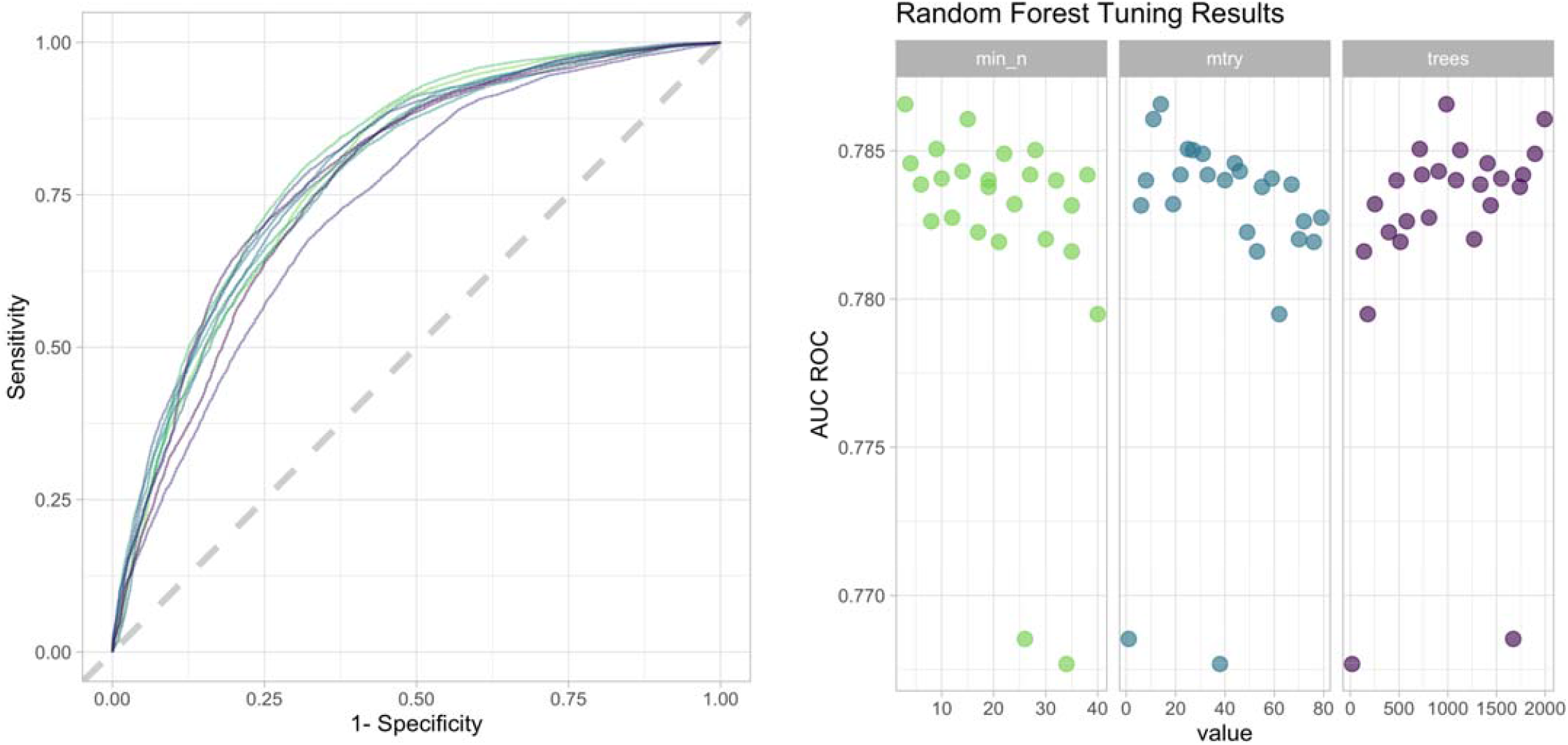
A) AUROC of the RF model used to calculate variable importance. B) The three hyperparameters used to tune the RF model that was used to calculate variable importance.

## References

1. Cohn BA, Cirillo PM, Murphy CC, Krigbaum NY, Wallace AW. SARS-CoV-2 vaccine protection and deaths among US veterans during 2021. Science (80-). 2022. doi:10.1126/science.abm0620

2. Eyre DW, Taylor D, Purver M, et al. Effect of Covid-19 Vaccination on Transmission of Alpha and Delta Variants. N Engl J Med. 2022. doi:10.1056/nejmoa2116597

3. Dexamethasone in Hospitalized Patients with Covid-19. N Engl J Med. 2021. doi:10.1056/nejmoa2021436

4. Emanuel EJ, Persad G, Upshur R, et al. Fair Allocation of Scarce Medical Resources in the Time of Covid-19. N Engl J Med. 2020. doi:10.1056/nejmsb2005114

5. Smith GB, Prytherch DR, Meredith P, Schmidt PE, Featherstone PI. The ability of the National Early Warning Score (NEWS) to discriminate patients at risk of early cardiac arrest, unanticipated intensive care unit admission, and death. Resuscitation. 2013. doi:10.1016/j.resuscitation.2012.12.016

6. Subbe CP, Kruger M, Rutherford P, Gemmel L. Validation of a modified early warning score in medical admissions. QJM - Mon J Assoc Physicians. 2001. doi:10.1093/qjmed/94.10.521

7. Hughes C, Pain C, Braithwaite J, Hillman K. “Between the flags”: Implementing a rapid response system at scale. BMJ Qual Saf. 2014. doi:10.1136/bmjqs-2014-002845

8. Seymour CW, Liu VX, Iwashyna TJ, et al. Assessment of clinical criteria for sepsis for the third international consensus definitions for sepsis and septic shock (sepsis-3). JAMA - J Am Med Assoc. 2016. doi:10.1001/jama.2016.0288

9. Liu VX, Lu Y, Carey KA, et al. Comparison of Early Warning Scoring Systems for Hospitalized Patients With and Without Infection at Risk for In-Hospital Mortality and Transfer to the Intensive Care Unit. JAMA Netw open. 2020. doi:10.1001/jamanetworkopen.2020.5191

10. Kar S, Chawla R, Haranath SP, et al. Multivariable mortality risk prediction using machine learning for COVID-19 patients at admission (AICOVID). Sci Rep. 2021. doi:10.1038/s41598-021-92146-7

11. An C, Lim H, Kim DW, Chang JH, Choi YJ, Kim SW. Machine learning prediction for mortality of patients diagnosed with COVID-19: a nationwide Korean cohort study. Sci Rep. 2020. doi:10.1038/s41598-020-75767-2

12. Moulaei K, Shanbehzadeh M, Mohammadi-Taghiabad Z, Kazemi-Arpanahi H. Comparing machine learning algorithms for predicting COVID-19 mortality. BMC Med Inform Decis Mak. 2022. doi:10.1186/s12911-021-01742-0

13. Shaw R, Lokshin AE, Miller MC, Messerlian-Lambert G, Moore RG. Stacking Machine Learning Algorithms for Biomarker-Based Preoperative Diagnosis of a Pelvic Mass. Cancers (Basel). 2022. doi:10.3390/cancers14051291

14. Džeroski S, Ž enko B. Is combining classifiers with stacking better than selecting the best one? Mach Learn. 2004. doi:10.1023/B:MACH.0000015881.36452.6e

15. Agius R, Brieghel C, Andersen MA, et al. Machine learning can identify newly diagnosed patients with CLL at high risk of infection. Nat Commun. 2020. doi:10.1038/s41467-019-14225-8

16. Charlson ME, Pompei P, Ales KL, MacKenzie CR. A new method of classifying prognostic comorbidity in longitudinal studies: Development and validation. J Chronic Dis. 1987. doi:10.1016/0021-9681(87)90171-8

17. Hastie T, Tibshirani R, Buja A. Flexible discriminant analysis by optimal scoring. J Am Stat Assoc. 1994. doi:10.1080/01621459.1994.10476866

18. Chen T, Guestrin C. XGBoost: A scalable tree boosting system. In: Proceedings of the ACM SIGKDD International Conference on Knowledge Discovery and Data Mining.; 2016. doi:10.1145/2939672.2939785

19. R Core Team. R: A language and environment for statistical computing. R Found Stat Comput. 2019.

20. Kuhn M, Wickham H. Tidymodels: a collection of packages for modeling and machine learning using tidyverse principles. 2020. https://www.tidymodels.org.

21. Wickham H, Averick M, Bryan J, et al. Welcome to the Tidyverse. J Open Source Softw. 2019. doi:10.21105/joss.01686

22. Couch S, Kuhn M. stacks: Tidy Model Stacking. 2020. https://cran.r-project.org/package=stacks.

23. Kuhn M. workflowsets: Create a Collection of “tidymodels” Workflows. 2021. https://cran.r-project.org/package=workflowsets.

24. Robin X, Turck N, Hainard A, et al. pROC: An open-source package for R and S+ to analyze and compare ROC curves. BMC Bioinformatics. 2011. doi:10.1186/1471-2105-12-77

25. Wickham H, Bryan J. readxl: Read Excel Files. 2019. https://cran.r-project.org/package=readxl.

26. Neuwirth E. RColorBrewer: ColorBrewer Palettes. 2014. https://cran.r-project.org/package=RColorBrewer.

27. Garnier S, Ross N, Rudis R, Camargo A, Sciaini M, Scherer C. Rvision - Colorblind-Friendly Color Maps for R. 2021. doi:10.5281/zenodo.4679424

28. Schloerke B, Cook D, Larmarange J, et al. GGally: Extension to “ggplot2.” 2020. https://cran.r-project.org/package=GGally.

29. Taiyun Wei, Simko V. R package “corrplot”: Visualization of a Correlation Matrix. 2017. https://github.com/taiyun/corrplot.

30. Kompaniyets L, Goodman AB, Belay B, et al. Body Mass Index and Risk for COVID-19–Related Hospitalization, Intensive Care Unit Admission, Invasive Mechanical Ventilation, and Death — United States, March–December 2020. MMWR Surveill Summ. 2021. doi:10.15585/mmwr.mm7010e4

31. Rajula HSR, Verlato G, Manchia M, Antonucci N, Fanos V. Comparison of conventional statistical methods with machine learning in medicine: Diagnosis, drug development, and treatment. Med. 2020. doi:10.3390/medicina56090455

32. Jassat W, Abdool Karim SS, Mudara C, et al. Clinical severity of COVID-19 in patients admitted to hospital during the omicron wave in South Africa: a retrospective observational study. Lancet Glob Heal. 2022;10(7):e961–e969. doi:10.1016/s2214-109x(22)00114-0

33. Ahnach M, Zbiri S, Nejjari S, Ousti F, Elkettani C. C-reactive protein as an early predictor of COVID-19 severity. J Med Biochem. 2020. doi:10.5937/jomb0-27554

34. Booth AL, Abels E, McCaffrey P. Development of a prognostic model for mortality in COVID-19 infection using machine learning. Mod Pathol. 2021. doi:10.1038/s41379-020-00700-x

35. Henry BM, Aggarwal G, Wong J, et al. Lactate dehydrogenase levels predict coronavirus disease 2019 (COVID-19) severity and mortality: A pooled analysis. Am J Emerg Med. 2020. doi:10.1016/j.ajem.2020.05.073

36. Nadim MK, Forni LG, Mehta RL, et al. COVID-19-associated acute kidney injury: consensus report of the 25th Acute Disease Quality Initiative (ADQI) Workgroup. Nat Rev Nephrol. 2020. doi:10.1038/s41581-020-00356-5

37. Goldstein JR, Lee RD. Demographic perspectives on the mortality of COVID-19 and other epidemics. Proc Natl Acad Sci U S A. 2020. doi:10.1073/pnas.2006392117

38. Lauring AS, Tenforde MW, Chappell JD, et al. Clinical severity of, and effectiveness of mRNA vaccines against, covid-19 from omicron, delta, and alpha SARS-CoV-2 variants in the United States: Prospective observational study. BMJ. 2022. doi:10.1136/bmj-2021-069761

39. Kipnis P, Turk BJ, Wulf DA, et al. Development and validation of an electronic medical record-based alert score for detection of inpatient deterioration outside the ICU. J Biomed Inform. 2016. doi:10.1016/j.jbi.2016.09.013

40. Liu VX, Bates DW, Wiens J, Shah NH. The number needed to benefit: estimating the value of predictive analytics in healthcare. J Am Med Informatics Assoc. 2019. doi:10.1093/jamia/ocz088

